# Fetal hemoglobin enables malaria parasite growth in sickle cells but augments production of transmission stage parasites

**DOI:** 10.1101/2024.09.05.24313101

**Authors:** Catherine Lavazec, Cheikh Loucoubar, Florian Dupuy, Jean-François Bureau, Isabelle Casadémont, Bronner P. Gonçalves, Swee Lay Thein, Mark Lathrop, Sandrine Laurance, Camille Roussel, Caroline Le Van Kim, Yves Colin, Mariane De Montalembert, Anavaj Sakuntabhai, Rick E. Paul

**Author notes:** Corresponding author Rick Paul; Institut Pasteur, Paris, France; **Email:**. C.L. & C.L. contributed equally.

## Abstract

Sickle cell trait is the quintessential example of the human response to malaria, providing protection against severe disease, but leading to sickle cell disease (SCD) in the homozygous state. Fetal Hemoglobin (HbF) reduces the pathology of SCD and several mutations lead to the prolonged production of HbF into childhood and adult life. HbF has been suggested to contribute to protection against malaria. Two long-term cohorts were genotyped for three quantitative trait loci associated with HbF production and analyzed for HbF titers, malaria clinical episodes and the production of parasite stages infectious to mosquitoes, gametocytes in asymptomatic infections. *Plasmodium falciparum* parasites were also grown *in vitro* in HbSS cells with measured levels of HbF. The genetic determinants of prolonged HbF production were associated with increased HbF titers and that increased HbF afforded protection from malaria disease but increased the production of gametocytes. The presence of HbF in sickled red cells was also shown in *in vitro* culture to enable parasite persistence in conditions otherwise deleterious for the parasite and enabled complete maturation of gametocytes. The beneficial personal effect of HbF, whether through protection against malaria or alleviating effects of SCD, is offset by increased parasite transmissibility and disease burden for the community. These individuals represent an important reservoir of infection and need to be targeted in elimination strategies.

**Key points:**

- Individuals with mutations associated with fetal hemoglobin production had fewer clinical episodes but produced more gametocytes.
- HbF in red blood cells enables gametocyte production at tissue level oxygen partial pressures at which normally parasite lysis would occur.

## Introduction

Transmission of the parasite from man to mosquito requires the presence of gametocyte stage parasites. Whilst there have been several epidemiological studies identifying risk factors for the presence of gametocytes, thereby offering a potential target group for treatment, none have been considered sufficiently robust.^1–3^ One notable feature was that the production of these gametocyte stages was found to have a strong human genetic basis (heritability), especially at sub-microscopic densities, but only during symptom-free infections.^4^ Mutations of the Beta-hemoglobin gene (HBB) have been shown to be associated with gametocyte carriage and transmission to mosquitoes,^2,5^ but explained little of this heritability.^4^

As encapsulated by the Red Queen hypothesis, hosts and pathogens are locked in a co-evolutionary arms race with adaptation and counter-adaptation to promote survival.^6^ The human genome has been thus remodeled over the millennia and hemoglobinopathies are quintessential examples of this positive selection, protecting from severe malaria.^7,8^ In turn, malaria parasites have been shown to be particularly efficient in responding to host defenses to ensure both their survival, through antigenic variation, and their transmission, through modulation of the sex ratio of the gametocytes, responsible for transmission to mosquitoes, thereby responding to the immediate human immunological and hematological response to infection.^9,10^

Hereditary persistence of fetal hemoglobin (HPFH) is a benign condition where the production of fetal hemoglobin (HbF) persists into adulthood and alleviates the severity of sickle cell disease and β-thalassemias.^11^ The protective effect afforded by these hemoglobinopathies against malaria has been suggested to be in part due to HbF.^12–14^ and gametocytes have been shown *in vitro* to be able to grow in fetal RBCs when added to cultures of normal HbA RBCs.^15^

Overall, this study aims to address the role that HbF plays not only in protecting from malaria but also in eliciting the parasite response to such an HbF rich environment in two long-term human cohorts in malaria endemic settings and the impact of HbF in sickle cells *in vitro*. Specifically the aims are to address: (1) the association of the three Quantitative Trait loci (QTL) previously shown to induce HbF production with HbF titers in the two study cohorts; (2) the association of these QTL with the number of clinical malaria episodes; (3) the association of these QTLs with gametocyte positivity in asymptomatic infections and (4) the impact of the presence of HbF in HbSS (sickle cells) RBCs in *in vitro* culture.

## Materials and Methods

### Study sites and populations

Two villages of differing malaria epidemiology were studied. Malaria transmission is perennial in Dielmo, where a river maintains larval breeding sites for the mosquitoes. By contrast, malaria transmission is strictly seasonal in Ndiop and dependent upon the rainy season that occurs from July-September. Such differing transmission has marked consequences on the epidemiology of malaria in the villages. This is most evident in the higher *P. falciparum* prevalence rates of infection in Dielmo (80+%) compared to the seasonal rates in Ndiop that change from 20% in the dry season to 70% in the rainy season.^16,17^ Peak incidence rates of clinical disease due to *P. falciparum* occur in very young children (2-5 years old) in Dielmo and then decline rapidly with age compared to 5-10 in Ndiop with only a gradual decrease with age.^18^ The acquisition of non-sterilising clinical immunity thus generates a much higher prevalence of asymptomatic infections in Dielmo. A field research station with a dispensary was built for the project in each village to identify all episodes of morbidity. Similar entomological surveys and identical strict clinical surveillance programs were carried out in both villages.

The family structure (pedigree) was available after a demographic census performed for every volunteer at his adhesion in the project. A verbal interview of mothers or key representatives of the household was used to obtain information on genetic relationships between studied individuals, their children, their parents, and to identify genetic links among the population. The total pedigrees, in Dielmo and Ndiop respectively, comprised 828 and 948 individuals, including absent or dead relatives, composed of 206 and 222 nuclear families (father – mother couples with at least one child) with averages of 3.6 and 3.8 children per family. In addition, previous typing with microsatellites has enabled the construction of a pedigree based on Identity-by-Descent using MERLIN.^19,20^

### Fetal Hemoglobin levels

Intra-venous blood samples (2ml) were taken from 497 individuals. The HbF proportion of total hemoglobin was measured by high pressure liquid chromatography (HPLC) on a BioRad Variant II system. We evaluated the heritability (*h*^2^) by using the SOLAR software package (version 2.1.4), as described previously.^4,21^

### Malaria related parameters measurement

As previously described,^20^ clinical case detection was both active and passive. A villager, who had volunteered to participate in the study, was visited daily at home by a physician, a technician, a nurse or medical field workers. Thick blood films were prepared and detailed medical examinations were made for villagers who had fever or any symptom that could be related to clinical malaria. A standardized questionnaire was completed for each episode of illness, recording clinical findings, treatment administered and response to treatment. None of the inhabitants received antimalarial chemoprophylaxis. The justification for not providing chemoprophylaxis within the project was based on the semi-immune status of the population, the level of chloroquine resistance in the area, and the permanent presence of a medical team in each village with daily home visits to each inhabitant. Urine tests were regularly carried out in the villagers to detect the presence of antimalarial drugs. Less than 2 percent of the results of tests carried out during the study period were compatible with unprescribed self-treatment. We considered an episode of *P. falciparum* clinical malaria in any case of fever (axillary temperature >= 37.5°C) or fever-related symptoms (headache, vomiting, subjective sensation of fever) associated with i) a *P. falciparum* parasite/leukocyte ratio higher than an age-dependent pyrogenic threshold previously identified in the patients from Dielmo or ii) a *P. falciparum* parasite/leukocyte ratio higher than 0.3 parasite/leukocyte in Ndiop.

The survey periods were defined by trimester for each inhabitant. Only person-trimesters with more than 30 days under survey in the village were included in the analysis. Person-trimesters with pregnancy during the trimester were excluded.

### Genotyping

***HBB* mutation:** the HbS mutation was genotyped by PCR (Polymerase Chain Reaction) - RFLP (Restriction Fragment Length Polymorphism). Following DNA extraction, a 559 bp fragment covering codon 6 of the β–globin gene (*HBB*) gene was amplified by PCR using the primers HbS_F : 5’-AGGGGAAAGAAAACATCAAGGGTC-3’ and HbS_R : 5’-ATAAGTCAGGGCAGAGCCATCTAT-3’. The amplification reaction was carried out using 5 µL of DNA in a reaction volume of 15 µL composed of MgCl_2_ [2.5mM], dNTPs [1mM], each primer [1µM], 1.5 µL PCR buffer (Qbiogene) and 0.04 µL Taq (Qbiogene). Amplification cycle conditions were: 4min at 94°C, and then 35 cycles of 30sec at 94°C, 30sec at 65°C, and 30sec at 72°C, with a final extension phase of 10 min. at 72°C. The amplified fragment (5 µL of DNA) was then digested by restriction enzyme Dde I (2U) in a reaction volume of 15 µL containing 1.5 µL 10x Buffer. Wildtype *HBB* yields 6 fragments of <u>201 + 97</u> + 89 + 88 + 50 + 37 base pairs, whereas HbS mutation yields 5 fragments of <u>298</u> + 89 + 88 + 50 + 37 bp.

Three QTL are associated with HPFH previously associated with the production of HbF on chromosomes 2, 6 and 11 (Figure S1 and Table S1).^22,23^ The following SNPs were genotyped to cover the linkage regions in the two QTL on chromosomes 2 & 6, as well as the Xmn-1 mutation on chromosome 11. SNPs were selected based on the minor allele frequencies in Yoruba populations in HapMap.

Chromosome 2: rs1427407, rs243027, rs6732518

Chromosome 6: rs11759553, rs11154792, rs9399137, rs6904897, rs4895441, rs1320963

Chromosome 11: Xmn-1

Their chromosomal positions and genotype frequencies are shown in Table S1.

### TaqMan Assay for SNP Genotyping

The PCR reaction was carried out in a 5 μl reaction containing 5 μl of PCR solution and 1 μl of DNA 1 ng/1 μl, which dry at 37°C before adding PCR solution. TaqMan Universal PCR Master Mix was used. PCR reaction mix consisted of 0.5 μl of 10X PCR buffer, 0.5 μl of 50mM MgCl_2_, 0.1 μl of Reference Dye, 0.02 μl of Platinum Taq (5U/μl) (Invitrogen), 0.312 μl of dNTPs 8 mM, 0.125 μl of 40X TaqMan probe, which consists of forward primer, reverse primer, and 2 fluorescence labeled probes, and 3.568 µl of H_2_O. After the ABI PRISM 96 well Optical Reaction plate was covered with ABI PRISM Optical Adhesive Covers, it was incubated at 95°C for 10 min, amplification was carried out for 40 cycles with the following temperature cycling parameters; 92°C for 0.15 min and 60°C for 1 min.

### *In vitro* experiments

#### Red blood cells

Hemoglobin genotypes were determined as above. The HbF proportion of total hemoglobin was measured by high pressure liquid chromatography (HPLC) on a BioRad Variant II system or by flow cytometry with anti-Human Fetal hemoglobin monoclonal antibody APC conjugated (Invitrogen) using a BD Accuri C6 cytometer. Erythrocytes were stored at 4°C and used within one week after collection. Erythrocytes were washed three times in RPMI 1640 before use.

#### Parasite culture

The *P. falciparum* clonal line B10 and the Pfs16-GFP line expressing GFP under the control of the gametocyte-specific promoter *Pfs16* were cultivated *in vitro* in human erythrocytes using RPMI 1640 medium supplemented with 10 % heat inactivated human serum, 100 µM hypoxanthine and 20 µg/ml gentamycin.^24,25^ For each experiment, synchronized schizonts obtained via magnetic purification were used to invade HbSS or HbAA erythrocytes at 5 % hematocrit. Cultures were maintained at 37°C in a gas environment of 1 - 2 % O2 / 5 % CO2 / 93 - 94 % N2. Gametocytemia and total parasitemia were daily monitored by Giemsa-stained blood smears and by flow cytometry analysis using a BD LSRII cytometer after DNA staining with Hoechst 33342 at 1:20000 (Life technologies).

#### Detection of parasites in HbF-containing cells by flow cytometry

HbSS or HbAA erythrocytes infected with the Pfs16-GFP line were fixed for 15 min at room temperature in 1X PBS / 2.7 % formaldehyde / 0.1 % glutaraldehyde and then permeabilized with 1X PBS / 1 % Octyl β-D-glucopyranoside (Sigma-Aldrich) for 15 min at room temperature. Cells were washed in 1X PBS, incubated in saturation solution (1X PBS / 1 % BSA / 2 % goat serum) for 15 min at room temperature and then incubated with anti-Human Fetal hemoglobin monoclonal antibody APC conjugated (Invitrogen) at 1:20 in saturation solution for 20 min at room temperature. After two washes in 1X PBS, cells were resuspended in saturation solution and analyzed using a BD Accuri C6 cytometer.

#### Detection of parasites in HbF-containing cells by immunofluorescence microscopy

HbSS or HbAA infected with the Pfs16-GFP line were fixed for 15 min at room temperature in 1X PBS / 1 % paraformaldehyde and then permeabilized with 1X PBS / 0.1 % Triton-X100 (Sigma-Aldrich) for 10 min at room temperature. Samples were washed three times in 1X PBS, smeared on glass slides and dried. The non-specific binding sites were saturated for 2 h in 1X PBS / 2 % BSA and the smears were then incubated with anti-Human Fetal hemoglobin monoclonal antibody APC conjugated (Invitrogen) at 1:500 for 1 h 30. After three washes in 1X PBS / 2 % BSA, the smears were incubated with Alexa594 anti-mouse secondary antibodies at 1:2000 (Life Technologies) and Hoechst 33342 at 1:20000. Smears were observed at 100X magnification using a Leica DMi8 fluorescence microscope.

### Statistics

Data for each malaria-related phenotype were first analysed using multivariate regression analysis according to the distribution of the data. This method enabled us to take into account known confounding factors such as age, year of study and transmission intensity. The residual unexplained variation in each phenotype was then calculated from the difference between the observed and expected values and then used as the phenotype for further genetic analysis.

As previously described,^13^ for clinical malaria attacks, we considered the number of clinical malaria attacks recorded during June 1990 to May 1998 in Dielmo and during July 1993 to May 1999 in Ndiop. The median (range) number of clinical *P. falciparum* malaria attack/person was 1 (0-69) in Dielmo and 2 (0-37) in Ndiop. The residual number of clinical *P. falciparum* attacks was transformed from the number of clinical malaria attacks during person-trimesters separately for each village by Poisson regression models including the effect of age, level of transmission and year of survey. To take into account the malaria exposure dependent acquired immunity in Dielmo, the models also controlled the effect of the proportion of time spent in the village for each individual from his/her birth to the beginning of person-trimester survey period. This confounding factor had no significant effect in Ndiop.

For gametocyte prevalence in asymptomatic infections: as previously described,^4^ asymptomatic *P. falciparum* infections were identified through monthly systematic blood slides taken from participating individuals from 1990–1998 and 1993–1998 in Dielmo and Ndiop respectively. Samples from individuals who had had a symptomatic episode within the last 30 days were excluded. Parasite positivity was established as follows. Thick and thin blood films were prepared and stained by 3% Giemsa stain. Blood films were examined under an oil immersion objective at 1000 magnification by the trained laboratory technicians and 200 thick film fields were examined to count the number of asexual and gametocyte parasite stages. Parasite species were identified on thin films and asexual parasite densities (per mL) were calculated from thick film by establishing the ratio of parasites to white blood cells (WBC) and then multiplying the parasite count by 8,000, the average WBC count per mL of blood. Gametocyte positivity was defined as the proportion of parasite positive infections that also carried gametocytes. The duration of gametocyte carriage for a single infection in endemic settings can last up to 30 days. The longevity and infectivity of gametocytes have been shown to persist for 3 weeks following chloroquine treatment of clinical cases. To increase the probability that only independent symptomatic or asymptomatic episodes (of gametocyte production) from the same individual were considered, consecutive samples with blood-stage malaria parasites of the same species within 30 days were excluded. Mixed parasite species infections were also excluded. As there were repeated measures for the same individual, a Generalized Linear Mixed Model was fitted with individual person as a factor in the random model. A binomial error structure was implemented (thus a logistic regression). Explanatory factors included date, which was classified annually by semester, reflecting the transmission seasons. Additional factors were gender and age factored initially into eight groups (<1, 1–4, 5–9, 10–14, 15–24, 25–39, 40–59 and 60+ years of age). The residual variance not explained by these ‘‘environmental’’ factors was generated and used for genetic analyses. Because a non-normal error distribution was used, Pearson rather than standardized normal residuals were generated. All statistical analyses were performed in Genstat version 22.^26^

### Ethics

The project protocol and objectives were carefully explained to the assembled village populations and informed consent was individually obtained from all subjects either by signature or by thumbprint on a voluntary consent form written in both French and in Wolof, the local language. Consent was obtained in the presence of the school director, an independent witness. For very young children, parents or designated tutors signed on their behalf. The protocol was approved by the Ethical Committee of the Pasteur Institute of Dakar and the Ministry of Health of Senegal. An agreement between Institut Pasteur de Dakar, Institut de Recherche pour le Développement and the Ministère de la Santé et de la Prévention of Senegal defines all research activities in the study cohorts. Each year, the project was re-examined by the Conseil de Perfectionnement de l’Institut Pasteur de Dakar and the assembled village population; informed consent was individually renewed from all subjects. Every person satisfying adhesion conditions could become a volunteer and every volunteer could leave the study at any time, therefore forming a dynamic open cohort. Further details of the study sites and adhesion criteria are previously described.^16,27^

#### Data sharing statement

Data set is available on Figshare with this private link: https://figshare.com/s/4af638e2e625e793c1dd

## Results

HbF levels from 497 (N=285 Dielmo, N=212 Ndiop) symptom-free individuals from two family-based cohorts in a malaria endemic zone in Senegal were measured and found to follow a log-series distribution (Figure S2), yielding a mean HbF level of 1.58% (variance 1.42%) in Dielmo and 1.39% (variance 1.00%) in Ndiop. HbF levels decreased exponentially with age (Figure S3).

Of 285 sampled individuals in Dielmo, 64% were blood slide positive for *Plasmodium falciparum* malaria parasites and 5.4% *P. falciparum* and *Plasmodium ovale* mixed infections. Only two *P. falciparum* blood slides had parasite densities in the range of 100-500 parasites/uL, the remainder had densities of less than 100 parasites/uL. All mixed infections had low densities (<100 parasites/uL) and were excluded from further analyses. In Ndiop, of 212 sampled individuals, 35% were *P. falciparum* positive. All parasite densities were less than 100 parasites/uL. Parasite infection was associated with increased HbF levels by 29.7% and 34.3% in Dielmo and Ndiop, respectively (Loglinear regression; P=0.033 & P=0.021). After accounting for the effects of age and parasite presence, gender not being found to be associated with HbF levels, the heritability of the residual HbF levels was estimated to be 43.1% (SE 10.9%, P = 4×10^-7^) in Dielmo and 48.7% (SE 16.2%, P= 4.8×10^-4^) in Ndiop.

The entire Dielmo cohort (597 individuals) were genotyped for the sickle mutation (*rs334*) and single nucleotide polymorphisms (SNPs) in the three QTL. Only two individuals were HbSS. Using the family-based likelihood ratio test (15), SNPs *rs1427407* on chromosome 2, *rs11759553* on chromosome 6 and the *Xmn1-HBG2* (*rs7482144*) in the HBB cluster were associated with HbF titre (P=3.2 x 10^-7^, P=0.0009 and P=0.017, respectively). Population based analyses confirmed the impact of *rs1427407* and *rs11759553*, accounting for 20% and 7% of variation in HbF levels, respectively. *Xmn1-HBG2* was found to have a significant effect on HbF only when co-occurring with HbS (HbSS and HbAS combined), accounting for 7% of variation in HbF levels.

HbF levels increased additively with increasing number of HbF-associated SNPs (N=308, 1 mutation, t=9.02, P<0.001; 2 or 3 mutations, t=12.2, P<0.001). We then examined the association of these HbF-related SNPs with the number of clinical malaria episodes and the proportion of asymptomatic infections that were positive for gametocytes. The number of clinical episodes decreased by 13% with inheritance of 1 and 40% with 2 or 3 HbF-related SNPs (N=313, 1 mutation t=1.15, P=0.25; 2 mutations t=2.96, P<0.05). This protective effect remained after exclusion of any individuals with HbS (N=283, 1 mutation t=1.31, P=0.19; 2 mutations, t=2.22, P<0.05). By contrast, gametocyte positivity increased with inheritance of any HbF-related SNPs by 27% (N=150: 1 mutation t=2.23, P<0.05; 2 mutations t=1.73, P=0.09). This association became even stronger upon exclusion of any individuals with HbS (N=137, 1 mutation t=3.85, P<0.001; 2 mutations, t=2.97, P<0.01) (Fig. 1).

**Figure 1.**
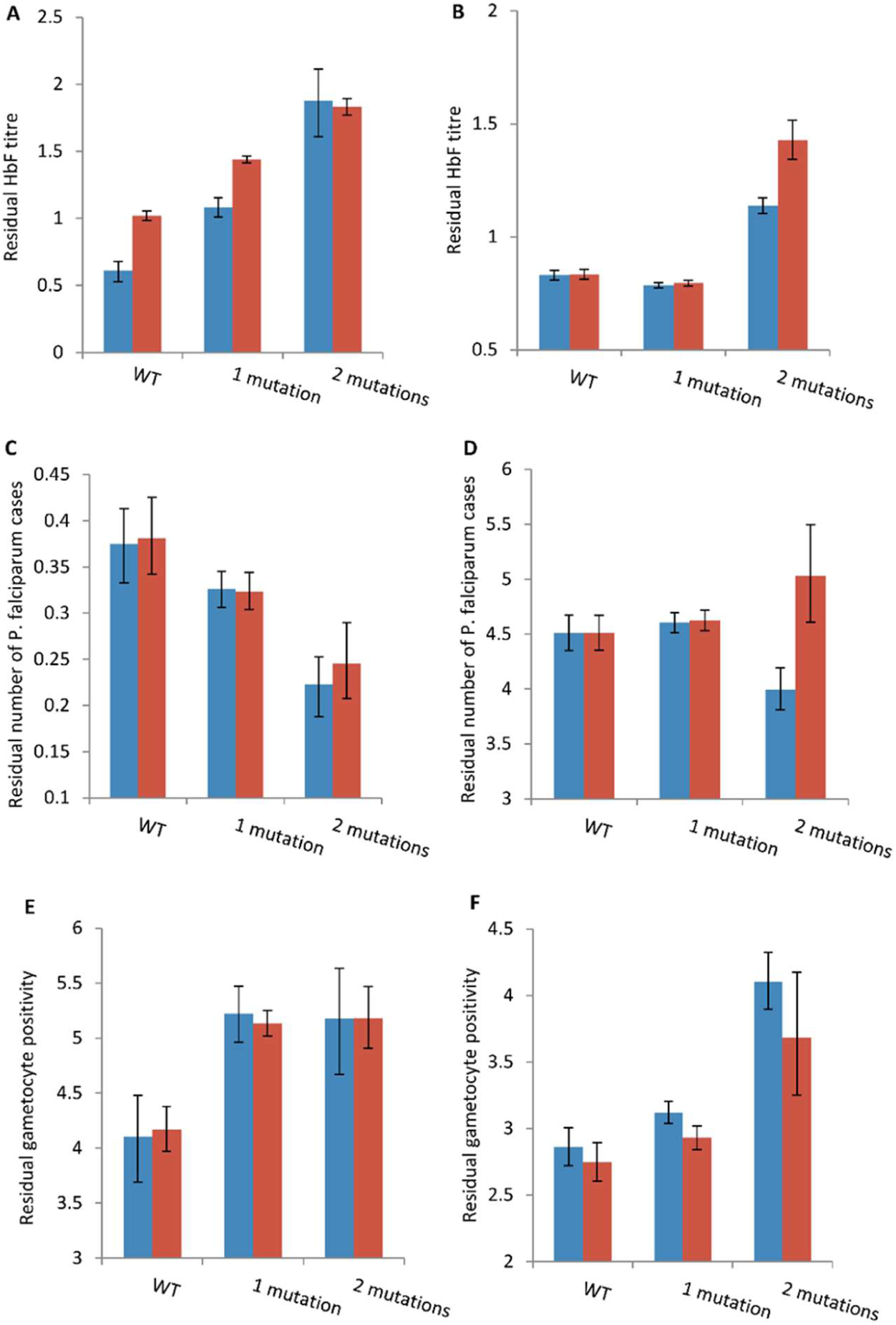
The effect of number of fetal hemoglobin mutations on HbF titre in (A) Dielmo and (B) Ndiop, Number of *P. falciparum* clinical episodes (C) Dielmo and (D) Ndiop and the Gametocyte prevalence rate (E) Dielmo and (F) Ndiop. WT – Wild Type. Shown are the means and standard of errors from the loglinear regression analyses on the residual values for each of three variables (see Supplementary materials). Blue histograms include individuals with sickle cell trait (HbS), red histograms have excluded the HbS individuals prior to the analyses.

We then replicated these findings in Ndiop (N=667) that had a ten-fold lower level of malaria transmission and composed of a different ethnicity. HbF was again found to increase additively with increasing number of HbF-related SNPs (N=124, 1 mutation, t=1.84, P=0.07; 2 or 3 mutations, t=7.91, P<0.001). The number of clinical malaria episodes again decreased with an increasing number of SNPs (N=389, 1 mutation t=0.51, P=0.61; 2 mutations t=2.02, P<0.05). This protective effect was lost after exclusion of any individuals with HbS, potentially because of simultaneous exclusion of *Xmn1-HBG2* (N=343, 1 mutation t=0.61, P=0.54; 2 mutations, t=1.16, P=0.24). By contrast, gametocyte positivity increased with the number of SNPs (N=229: 1 mutation t=1.53, P=0.13; 2 mutations t=5.01, P<0.001) and this association was maintained even upon exclusion of any individuals with HbS (N=203, 1 mutation t=1.07, P=0.29; 2 mutations, t=2.16, P<0.05) (Fig. 1).

We confirmed that *rs1427407, rs11759553 and Xmn1-HBG2* (*rs7482144*) were associated with increased production of HbF and showed that HbF-related SNPs protect from clinical malaria, but increase gametocyte prevalence and thus parasite transmissibility in two sites differing ten-fold in malaria transmission intensity.

To explore how HbF influences parasite growth, we took the extreme case of growth within HbSS erythrocytes. Early work showed that cultures of *P. falciparum* in HbSS erythrocytes under low oxygen (1-5%) exhibit rapid inhibition of growth.^28,29^ The sickling of HbSS erythrocytes kills and lyses most or all of the intracellular parasites in less than 6 hours, leading to the death of all parasites in 4 days.^28,30^ Here, we show that presence of HbF in HbSS erythrocytes allows the replication and development of *P. falciparum* asexual parasites during at least 8 days (Figs. 2A-B). HbF retards the polymerization of deoxy-sickle hemoglobin, the root cause of sickle cell disease, thereby enabling parasite development (Figs 2C-D).^31^ Under these conditions, although the parasite replicates at a lower rate than in normal HbAA cells, RBCs containing HbF allow appearance and complete maturation of gametocytes (Figs. 2C-F).

**Figure 2.**
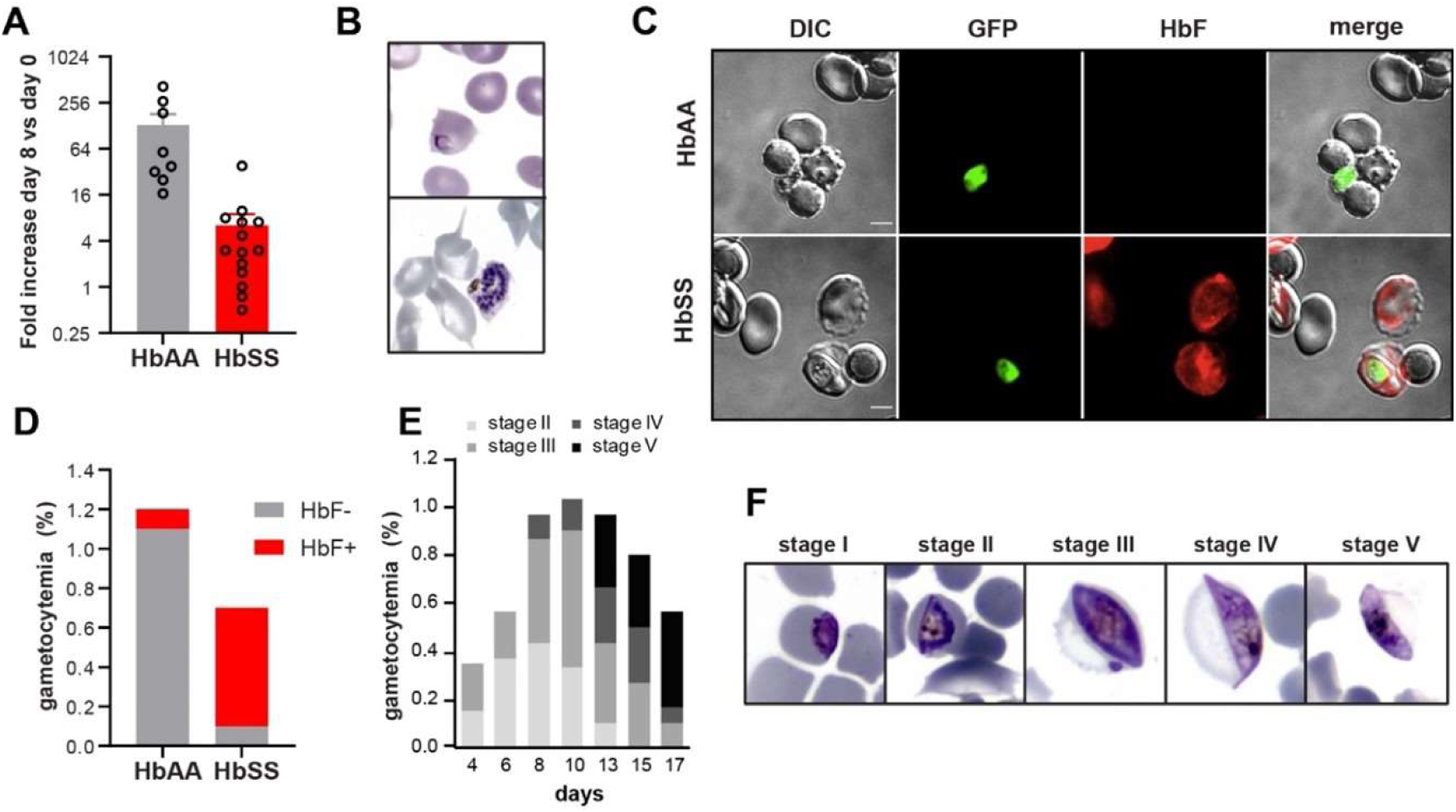
Development of *P. falciparum* in HbSS erythrocytes containing HbF. **A**. Increase in asexual parasitemia after 8 days of culture in HbAA or HbSS erythrocytes under an atmosphere of 1% to 2% O_2_. Circles show the number of independent experiments in blood from 8 HbAA donors and 14 HbSS donors containing HbF (from 2.1 to 21%). Parasitemia increased in blood of 11 out of 14 HbSS donors. **B**. Giemsa-stained images of ring-infected (top panel) or schizont-infected (bottom panel) or HbSS erythrocytes containing HbF. **C**, **D**. The presence of GFP-expressing gametocytes (green) in HbAA or HbSS erythrocytes containing 56% HbF-positive cells was analyzed by immunofluorescence analysis (**C**) and by flow cytometry (**D**). Erythrocyte staining with fetal hemoglobin monoclonal antibody highlighted that gametocytes preferentially develop in HbF-positive cells. DIC: differential interference contrast. The bars represent 2 µm. **E**, **F**. Follow-up of gametocytemia on Giemsa-stained smears show that gametocytes are able to complete their maturation in HbSS erythrocytes containing 12% HbF. Cultures were grown during 17 days under an atmosphere of 5% CO_2_, 1% O_2_ and 94% N_2_.

## Discussion

The occurrence of several QTL that are associated with hereditary persistence of fetal hemoglobin (HPFH) and found at high frequencies underlines the importance of HbF in alleviating the negative effects of hemoglobinopathies. Our studies show that these HbF QTL also contribute to protection from clinical malaria, concurring with previous *in vitro* and mouse model studies demonstrating reduced parasite replication in the presence of HbF.^13,32^ The strong effect of these QTL, especially *rs1427407*, on gametocyte production is consistent with *in vitro* studies demonstrating an induction of gametocyte production in reticulocytes that were derived from HPFH and sickle cell patients.^33^ Gametocyte production has been associated with the hematological environment: abnormal hemoglobin genes (S and C) and anemia induce gametocyte production *in vivo*,^1,4,5,34^ suggesting a response by the parasite to the prevailing hematological condition.

Dysregulated expression of the oncogene *BCL11A*, wherein lies *rs1427407* (erythroid-specific enhancer),^35^ may influence F cell production by affecting the kinetics of erythropoiesis.^11^ Stress erythropoiesis, which is associated with increased HbF, implicates erythropoietin in initiating downstream signaling pathways that activate *HBG* expression.^36^ The *BCL11A*-associated SNP reduces its expression and disrupts binding of transcription factors crucial for switching from gamma to beta globin production.^37,38^ The absence of these transcription factors enables binding of TGF-beta-induced Kruppel-like factors, KLF10 and KLF11, with subsequent increase in gamma globin production.^38^ High levels of TGF-beta are associated with an anti-inflammatory response to parasite infection and hence predict a positive association of parasitic infection and HbF levels in individuals carrying the *BCL11A*-associated SNP, as observed here. Our previous genome linkage analyses on asymptomatic parasite infections in these populations suggested the locus containing *KLF11* (LOD score ≈ 2.5) (Figure S4), consistent with the result observed here.^20^

The high frequency of HbF QTL in sickle cell and beta-thalassemia conditions, both of which confer resistance to malaria, likely provides a strong signal for parasite sexual differentiation. Hence, whilst the individual remains protected from the severe consequences of infection, gametocyte survival in HbF RBCs will increase malaria transmission at the community level. Thus, once again the parasite has developed a remarkable capacity to respond productively to an environment hostile to its proliferation. Humans, on the other hand, have accumulated successive mutations to alleviate the negative effects of previous malaria protective mutations, but in doing so ironically enable parasite survival.

## Acknowledgments

We are grateful to the villagers of Dielmo and Ndiop for their participation and continued collaboration and to the field workers for their active contribution in this project. We would like to thank Laetitia Claer for her advice and support. This work was supported by Institut Pasteur, Paris, Institut Pasteur de Dakar, CNRS, INSERM and the Laboratoire d’Excellence GR-Ex (ANR-11-LABX-0051).

## Author Contributions

Catherine Lavazec: Conceptualization, Experimentation, Writing. Cheikh Loucoubar: Data analyses, Writing. Florian Dupuy: Experimentation. Jean-François Bureau: Molecular analyses, Writing. Isabelle Casadémont: Molecular analyses. Bronner Gonçalves: Molecular analyses, Writing. Swee Lay Thein: Conceptualization, Writing-Reviewing and Editing. Mark Lathrop: Conceptualization, Writing-Reviewing and Editing. Camille Roussel: Experimentation. Sandrine Laurance: Provided input, Writing-Reviewing. Caroline Le Van Kim: Provided input, Writing-Reviewing. Yves Colin: Provided input, Writing-Reviewing. Mariane De Montalembert: Conceptualization, Patient inclusion. Anavaj Sakuntabhai: Conceptualization, Writing-Reviewing and Editing. Richard Paul: Molecular and data analyses, Literature review, Writing.

## Disclosure of conflicts of interests

Authors declare no competing interests.

## Supporting Information

**Figure S1.**
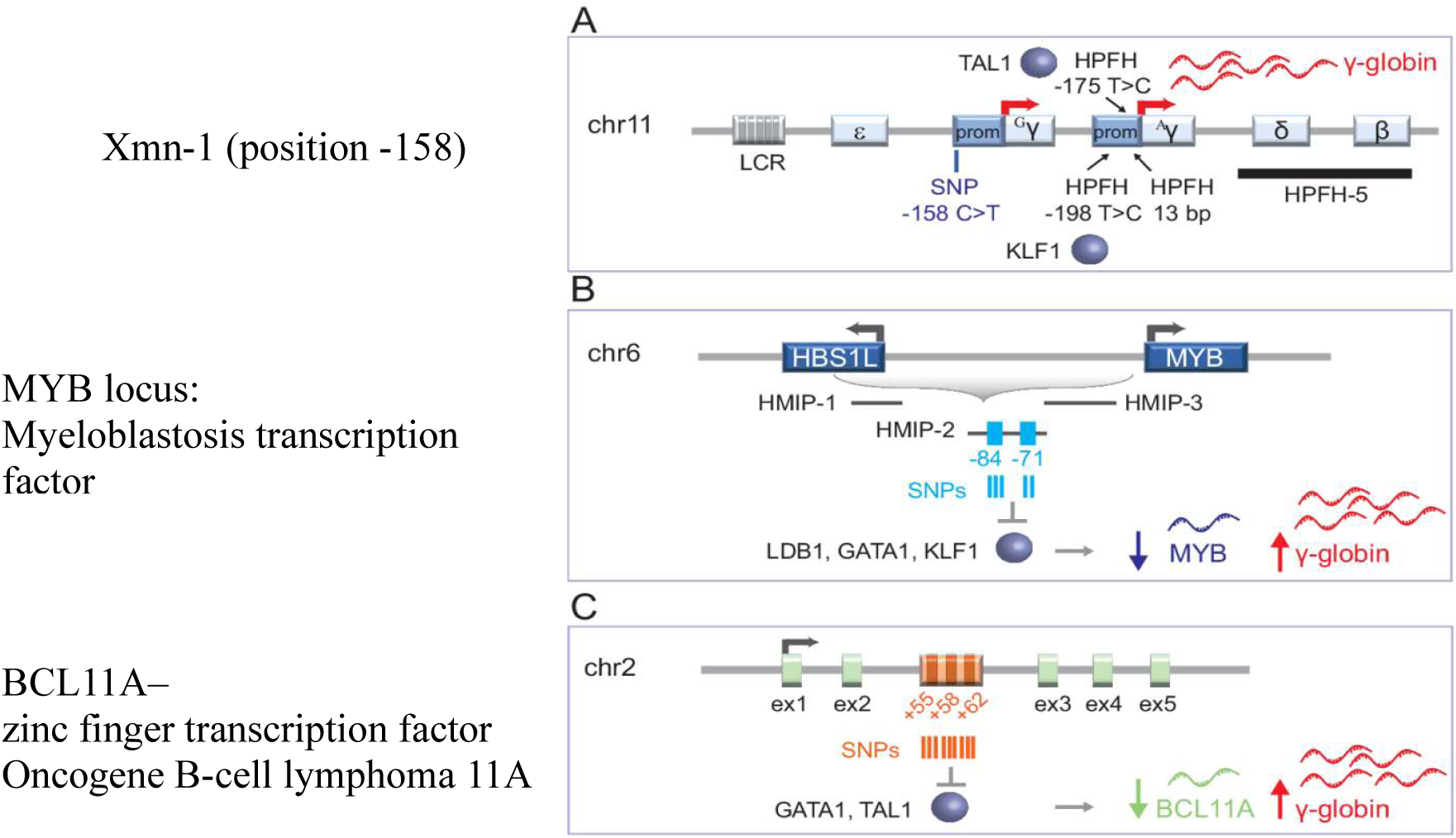
Three QTLs associated with HPFH (Image from (27))

**Figure S2.**
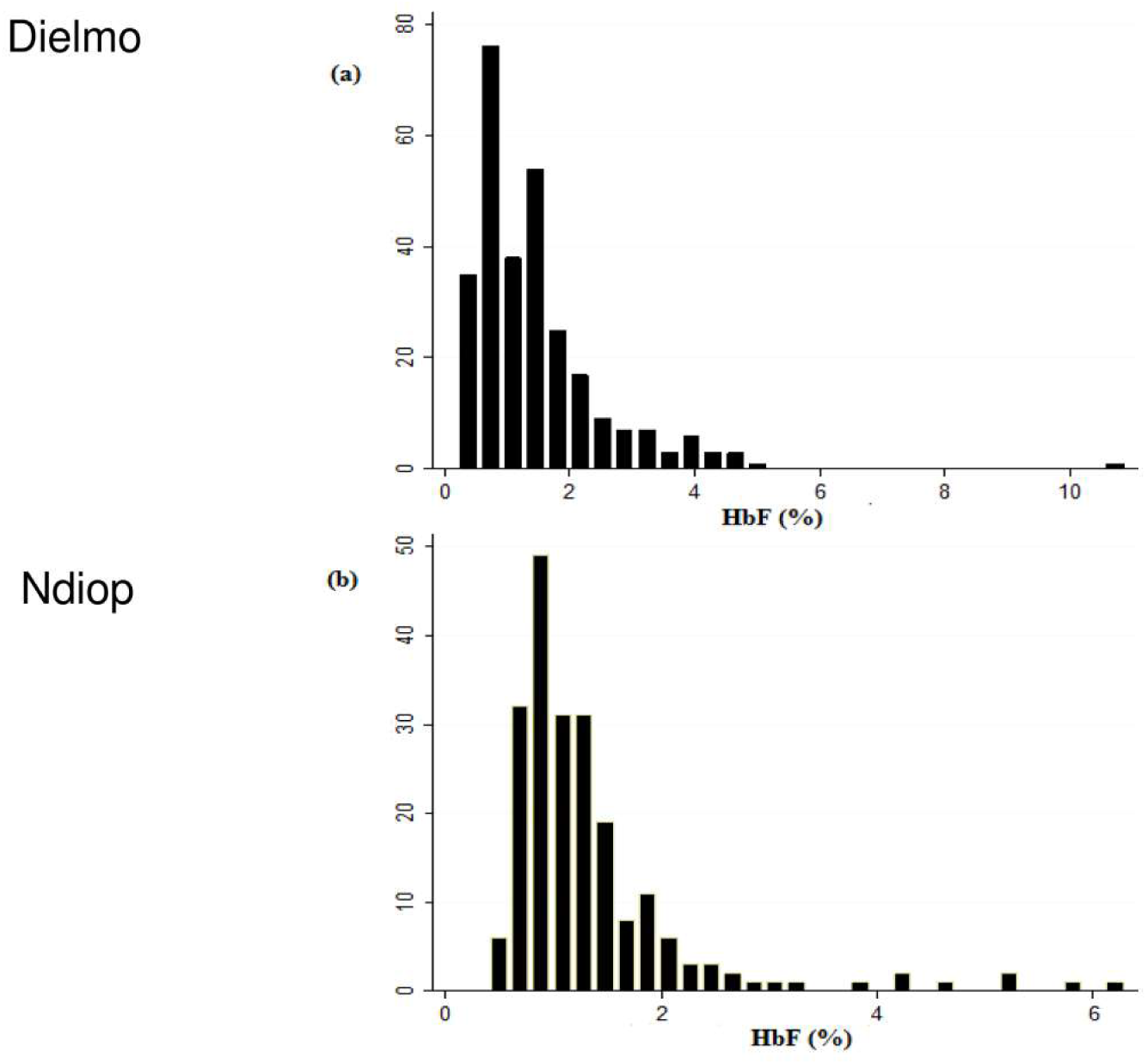
Distribution of HbF levels in the two cohorts.

**Figure S3.**
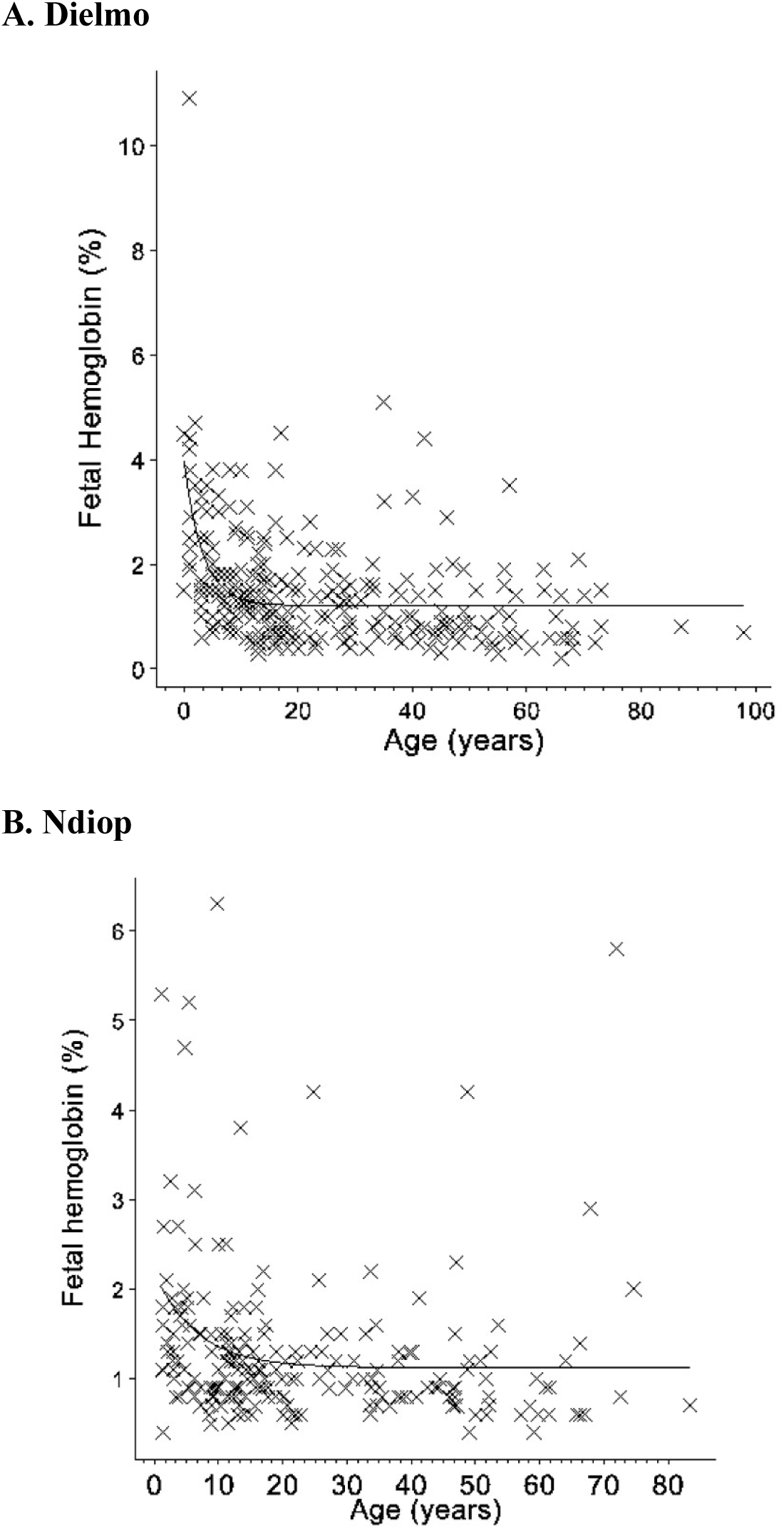
Decrease of HbF levels as a function of age in the two cohorts.

**Figure S4.**
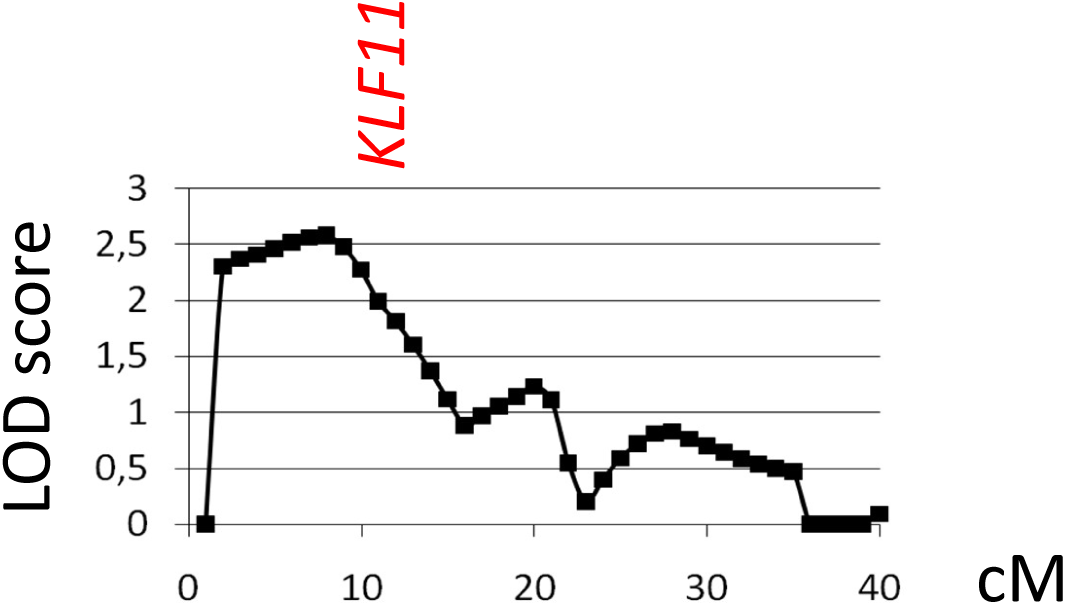
Genome scan linkage analysis results of the prevalence of asymptomatic infections in individuals from the Dielmo cohort (25). Shown is the result from Chromosome 2p and the location of the *KLF11* gene.

**Table S1.** Genotype frequencies of selected mutations.

| Chromosome | Position | SNP | Genotype | Dielmo | Ndiop |
| --- | --- | --- | --- | --- | --- |
| 11 | Beta-globin gene | HBB | AA | 352 | 456 |
|  |  |  | AS | 37 | 60 |
|  |  |  | SS | 2 | 0 |
| 11 | -158 of HBG2 gene | Xmn1 | 22 | 216 | 255 |
|  |  |  | 24 | 148 | 196 |
|  |  |  | 44 | 20 | 31 |
| 2 | 60,379,872 | rs243027 | 33 | 28 | 23 |
|  |  |  | 34 | 120 | 134 |
|  |  |  | 44 | 231 | 312 |
| 2 | 60,481,462 | rs6732518 | 22 | 57 | 54 |
|  |  |  | 24 | 196 | 215 |
|  |  |  | 44 | 119 | 191 |
| 2 | 60,490908 | rs1427407 | 33 | 168 | 239 |
|  |  |  | 34 | 149 | 188 |
|  |  |  | 44 | 47 | 38 |
| 6 | 135,061,842 | rs6904897 | 33 | 46 | 38 |
|  |  |  | 34 | 185 | 169 |
|  |  |  | 44 | 148 | 259 |
| 6 | 135,097,880 | rs9399137 | 24 | 16 | 31 |
|  |  |  | 44 | 362 | 628 |
| 6 | 135,101,158 | rs11759553 | 11 | 89 | 105 |
|  |  |  | 14 | 185 | 220 |
|  |  |  | 44 | 104 | 133 |
| 6 | 135,105,435 | rs4895441 | 11 | 340 | 417 |
|  |  |  | 13 | 36 | 94 |
| 6 | 135,110,502 | rs11154792 | 22 | 12 | 16 |
|  |  |  | 24 | 63 | 99 |
|  |  |  | 44 | 253 | 319 |
| 6 | 135,122,074 | rs1320963 | 11 | 54 | 63 |
|  |  |  | 13 | 153 | 226 |
|  |  |  | 33 | 127 | 165 |

## Notes

### Competing Interest Statement

The authors have declared no competing interest.

### Funding Statement

The authors acknowledge the financial support from the Institut Pasteur, Paris, Institut Pasteur de Dakar, CNRS, INSERM and the Laboratoire Excellence GR-Ex (ANR-11-LABX-0051).

### Author Declarations

Ethics committee of the Minstry of Health Senegal gave ethical approval for this work

